# Congenital syphilis in Argentina: experience in a pediatric hospital

**DOI:** 10.1101/2020.06.16.20132795

**Authors:** Luciana Noemí Garcia, Alejandra Destito Solján, Nicolas Falk, Nicolás Leonel Gonzalez, Griselda Ballering, Facundo García Bournissen, Guillermo Moscatelli, Samanta Moroni, Jaime M Altcheh

**Author notes:** Corresponding author: MD, PhD. Luciana N García, Servicio Parasitología- Chagas. Hospital de Niños Ricardo Gutierrez. Gallo 1330, C1425EFD, Buenos Aires, Argentina Tel/FAX: 54 011. 4964-4122. These authors contributed equally to this work (Luciana Noemí Garcia, Alejandra Destito Solján).

## Abstract

Although congenital syphilis (CS) is preventable, it is still an important health problem worldwide. Recently, an increase in the number of primary and congenital syphilis cases has been observed. Fetal infection can be particularly aggressive, but newborns can be asymptomatic at birth and run the risk of developing systemic compromise with a poor prognosis.

We conducted a study (1987-2019) analyzing the medical records of CS diagnosis cases assisted at the Buenos Aires Children’ Hospital. Sixty-one patients were included. Information about demographics, clinical and laboratory findings, *T. pallidum* serology and treatment was collected. Median age at diagnosis was 2 months (IQ 1-6 months). The distribution of cases showed a bimodal curve, with a peak in 1993 and in 2017. The main clinical findings were: bone alterations in 36/61 (59%); hepatosplenomegaly in 33/61 (54.1%); anemia in 32/51 (62.8%); skin lesions 26/61 (42.6%) and renal compromise in 15/45 (33.3%). Cerebrospinal fluid was studied in 50/61 (81.9%); 5 (10%) were abnormal (reactive VDRL and/or cell alteration count). Only 23 (60.5%) patients had nontreponemal titers fourfold higher than their mothers did. Intravenous penicillin G for an average of 10-14 days was prescribed in 60/61 subjects and one patient received ceftriaxone. Remarkably, only 28 (46%) mothers were tested for syphilis during pregnancy.

During follow-up, a decrease in RPR titers was observed reaching seroconversion in 31/34 (91%) subjects at a median of 19.2 months after treatment. Treponemal titers (TPHA) remained reactive.

Our results highlighted that an increase in the number of cases of CS is occurring in our population with high morbidity related to delayed diagnosis. A good therapeutic response was observed.

CS requires a greater effort from obstetricians to adequately screen for the disease during pregnancy and pediatricians should be alert in order to detect cases earlier, to provide an adequate diagnosis and treatment of CS.

**Author Summary:** CS is caused by mother-to child transmission. Although screening of pregnant women and treatments are available, new cases are increasing worldwide. We reviewed the medical records of CS-patients assisted in our hospital over the past 30 years. Our results showed that there was an increase in the number of CS cases. At birth, most children were asymptomatic and later developed CS clinical manifestations. Penicillin treatment, and in one case ceftriaxone, was prescribed with a good clinical response. Nevertheless, one infant died, four had persistent kidney disorders and one showed bone sequelae damage. Spinal lumbar puncture did not modify therapeutic decisions. In the follow-up, a decrease in nontreponemal antibodies was observed as a marker of treatment response. We concluded that the detection and treatment of CS remains a great challenge for clinical practice in our region.

It is crucial that pediatricians and obstetricians give greater attention and make a greater effort to detect this neglected disease in an attempt to reverse its upwards trend.

## Introduction

Congenital syphilis (CS) is the result of a transplacental infection caused by *Treponema pallidum pallidum* (TPA) and affects vulnerable populations with social and economic problems [1]. In 2015, PAHO estimated that there were 22,800 cases of CS with a rate of 1.7 cases per 1,000 live births in Latin America and an incidence of 22,800 new cases in that year [2]. In Argentina the number of syphilis cases has doubled or even tripled in recent years as a result of an increase in primary syphilis in the population of childbearing age, which has produced an increase in the number of CS cases [3]. In this scenario, adequate screening and treatment of infected mothers is vital to avoid transplacental transmission of TPA. [1] [4]

Another window of opportunity for diagnosis of CS is the neonatal and infancy period. Nevertheless, since the diagnosis is based on serological studies the interpretation of results is complex and often confusing due to the passive transplacental transfer of maternal antibodies [4]. The majority of infected newborns are asymptomatic and, if they are nor diagnosed and treated, the TPA infection persists silently. The resultant inflammatory response continues in tissues and may develop manifestations of CS months or years later. [5].

Our main objective was to describe the medical experience relating to the detection, treatment, clinical and serological evolution of patients with CS assisted in our hospital during the last 30 years.

## Methods

A Prospective cohort study with retrospective data collection was conducted in a cohort of pediatric patients with congenital syphilis, between February 1987 and June 2019, assisted at the Servicio de Parasitología-Chagas, Hospital de Niños “Ricardo Gutierrez”, a tertiary care hospital without a maternity unit.

### Case definition criteria

A) Newborns or infants with reactive TPA serology, mother with syphilis during pregnancy and/or clinical evidence of CS
Exclusion criteria: patients with acquired syphilis.

Adequate syphilis screening during pregnancy was defined by at least one nontreponemal serological test at the first trimester and another at the third trimester. Demographics, clinical findings, TPA serology, general laboratory, complementary studies, and treatment prescribed data were collected.

The serological tests used were the nontreponemal test: Rapid plasma reagin (RPR) or Venereal Disease Research Laboratory (VDRL) and the treponemal test: *Treponema pallidum* haemagglutination (TPHA).

Maternal data were collected for serological pregnancy control and antibiotic treatment.

Descriptive statistics were used for the variables of interest. Continuous variables are presented as means with 95% CI or medians and interquartile range. Categorical variables are represented in percentages. The disappearance kinetics of serum antibodies were analyzed using survival analysis (Kaplan-Meyer). Analyses were performed with R software v3.0 (R Core Team 2018. R Foundation for Statistical Computing, Vienna, Austria. https://www.R-project.org/).

## Ethics Statement

The Ethics Committee and Review Board of the Hospital de Niños Ricardo Gutierrez approved this study (approval number: DI-2020-159-GCBA-HGNRG).The study is registered in clinical trials.gov (NCT04137601) and adhered to the tenets of the Declaration of Helsinki. As part of our regular clinical practice all patients sign a consent form to their data and/or images used for academic purposes anonymously. In the case of a patients’ clinical manifestation picture were performed, previous oral consent was obtained from the parents/guardian of the patients.

## Results

Out of 100 clinical charts analyzed, 61 patients fulfilled inclusion criteria for CS. The remaining 39 were patients with acquired syphilis. The data of complementary studies is summarized in a flowchart in Fig 1.

**Fig 1.**
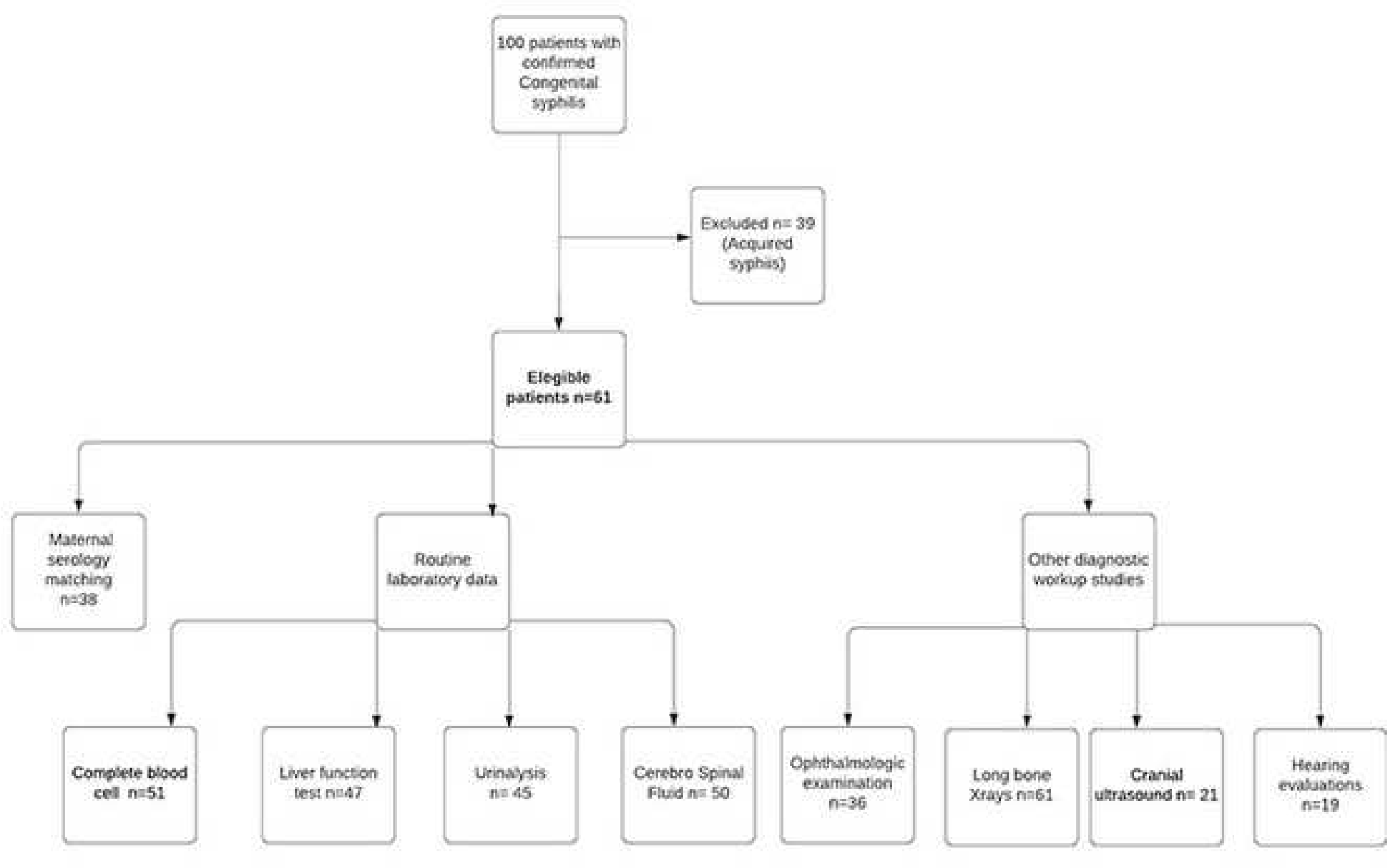
Flowchart of patient inclusion process and complementary studies.

A bimodal time curve was observed with a peak in 1993 and another in 2017 (Fig 2).

**Fig 2.**
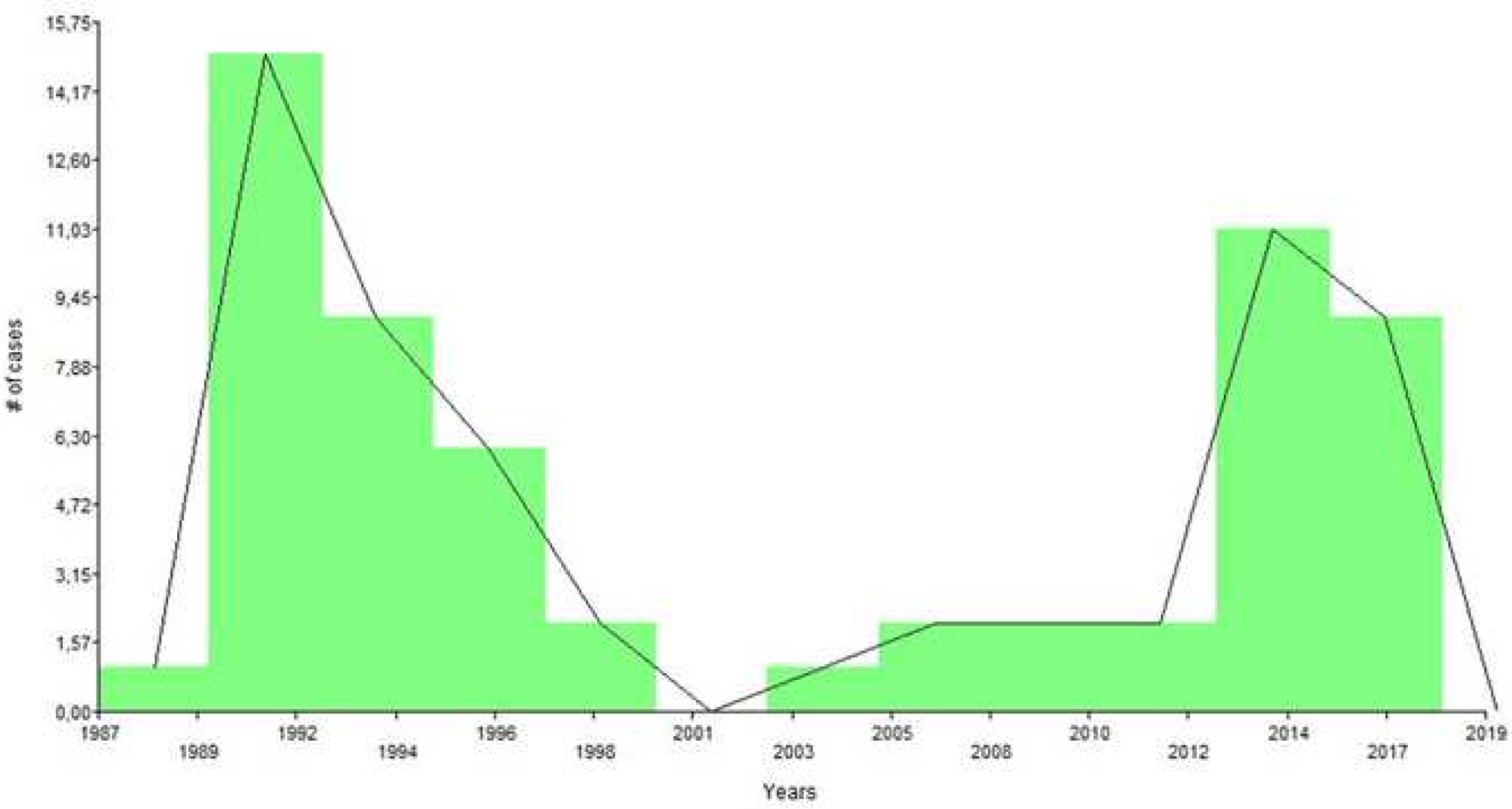
Number of congenital syphilis cases. Histogram shows registered cases by year.

As regards perinatal data, (Table 1), 20.6% of mothers were younger than 20 years of age. Adequate TPA serological control was carried out in only 27/57 (47%) women during pregnancy. Only 4 mothers received penicillin treatment. Three mothers received 3 doses of benzathine penicillin in the second trimester and the other one, received two regimens of 3 doses of penicillin ending within the last month of pregnancy. Serological data from 4 mothers were not available: 3 children were adopted and one mother died postpartum.

**Table 1.** Perinatal data.

| Perinatal data |  |  |
| --- | --- | --- |
| Pregnancy screening | n | % |
| Adequate | 27 | 44.3 |
| Uncontrolled pregnancy | 27 | 44.3 |
| Inadequate | 3 | 4.9 |
| No available data | 4 | 6.5 |
| **Birth outcome** | **n** | **%** |
| Preterm (< 37 weeks) | 15 | 24.6 |
| Low birth weight/ IUGR<sup>a</sup> | 15 | 24.6 |
| Full term babies | 31 | 50.8 |

<sup>a</sup> IUGR: Intrauterine Growth Restriction

The median age at diagnosis was 2 months (IQ25-75: 1-6 months), ranging from 1 day to 8.5 years. Early CS was diagnosed before one month of age in 6 patients, between one to six months of age in 40 patients, and between 1 to 2 years old in 9 patients. Late CS was diagnosed in 6 children older than 2 years (Fig 3). In 3 patients congenital coinfection was detected: cytomegalovirus in 2 patients and hepatitis B in 1. All patients showed negative HIV serology.

**Fig 3.**
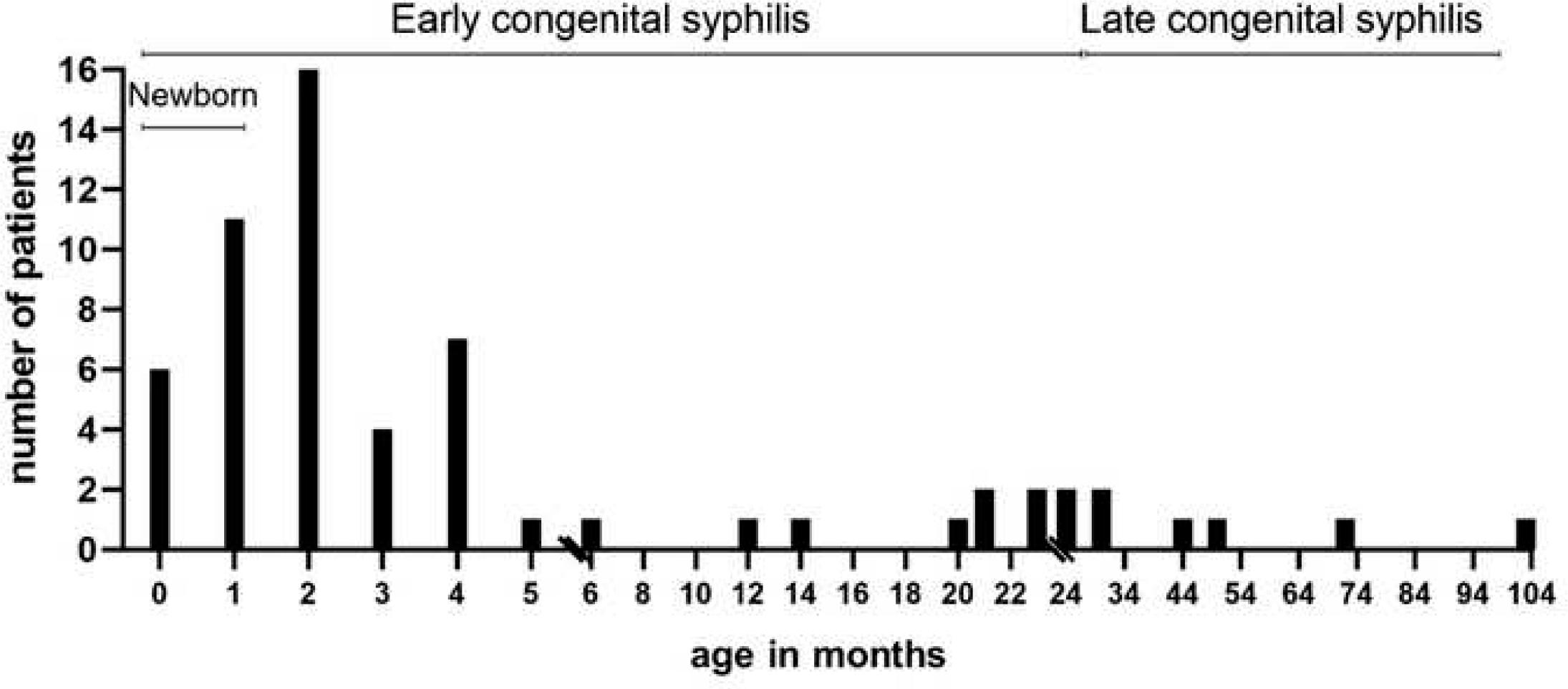
Timeline of the age at diagnosis. Bar chart shows the number of patients diagnosed at different ages and the period of early and late congenital syphilis.

At birth, 48/61 (78%) were asymptomatic. In the following weeks 40/48 developed symptoms of CS (media: 9.4, 95% CI:3.6-15.1 months) and 8 remained asymptomatic. These asymptomatic cases were diagnosed at different ages (media: 25.63, 95% CI:6.3-45) as follows: 2 cases were siblings of an index case, 2 cases by pre-surgical serological testing and 4 by serological screening due to the lack of information about TPA serology at birth.

In summary, at diagnosis, 53 (86.8%) CS infants were symptomatic and 8 (13.1%) asymptomatic (Fig 4).

**Fig 4.**
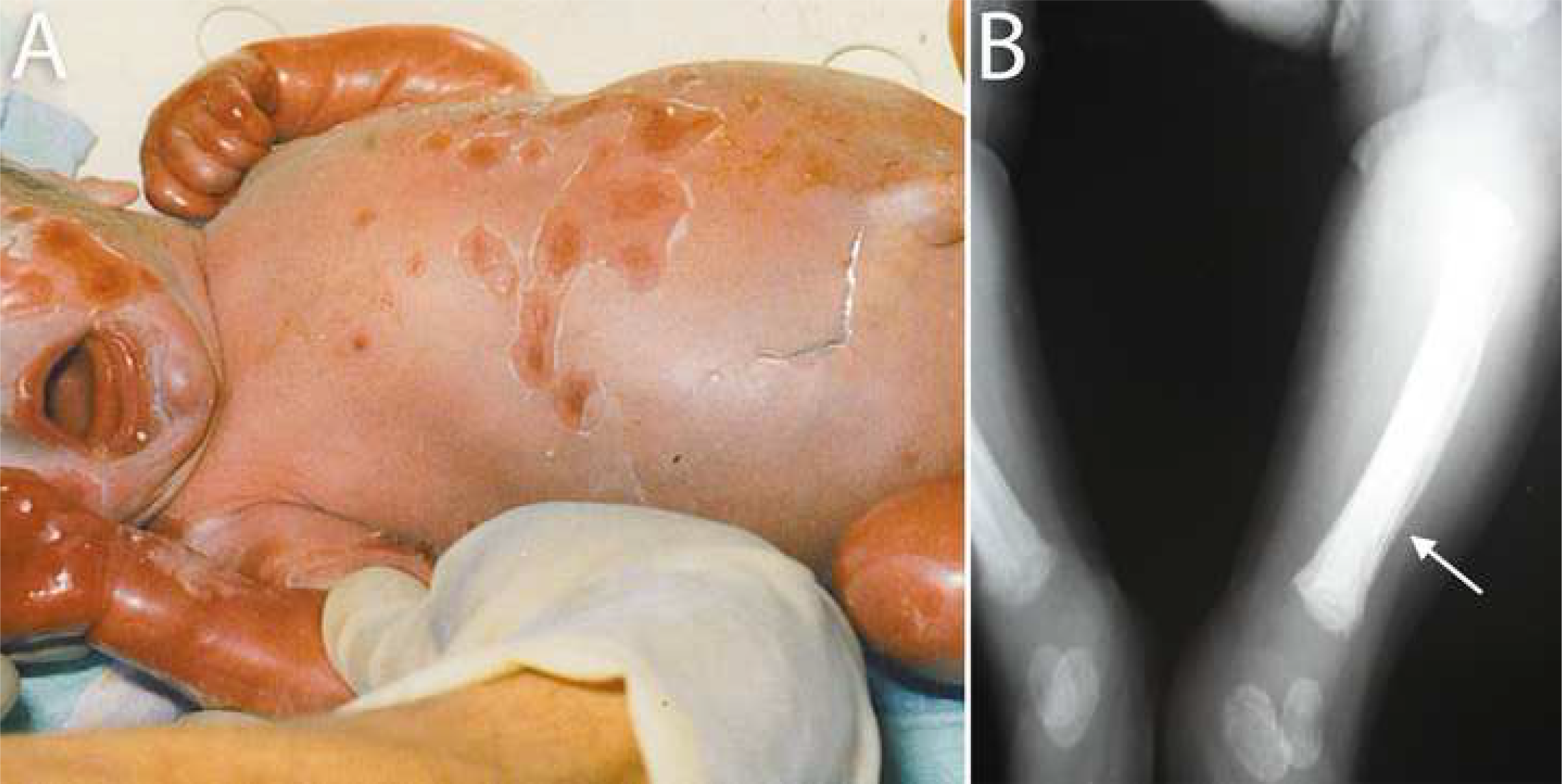
Symptomatic newborn. (A) Term male newborn, adequate for gestational age with widespread erythroderma, vesiculobullous eruptions and desquamation (pemphigus syphiliticus). (B) Bone involvement lesions in the same patients are showed: periosteal detachment (arrow).

Demographic data as well as clinical findings at diagnosis are presented in Table 2. Our hospital is a pediatric referral center without a maternity unit. The analysis of the place of residence showed that only 11 (18%) lived in Buenos Aires City, 41(67.3%) in nearby cities and 3 (5%) came from neighboring countries. In 6 (9.8%) cases data was not available. Low socioeconomic resources and limited access to the health system were registered in the assisted population.

**Table 2.** Demographic and clinical findings at diagnosis.

| **Age** |  |  |
| --- | --- | --- |
| Median | 2 months |  |
| Range | 1 day- 8.5 years |  |
| IQ 25-75% | 1-6 months |  |
| **Sex** | **n** | **%** |
| Female | 29 | 47.5 |
| Male | 32 | 52.5 |
| **Clinical status at diagnosis (n=61)** | **n** | **%** |
| Symptomatic | 53 | 86.8 |
| Asymptomatic | 8 | 13.1 |
| **Clinical findings <sup>a</sup>** | **n** | **%** |
| Bone involvement | 36 | 59 |
| Periostalgia | 36 | 100 |
| Pseudoparalysis | 13 | 36 |
| Saddle nose | 1 | 2.7 |
| Saber shins | 2 | 5.5 |
| Hepatosplenomegaly | 33 | 54.1 |
| Dermatological manifestations | 31 | 50.8 |
| Maculopapular rash | 17 | 27.8 |
| Palms and soles' rash | 17 | 27.9 |
| Pemphigus | 15 | 24.6 |
| Desquamation lesions | 11 | 18 |
| Jaundice | 11 | 18 |
| Condyloma lata | 2 | 3.3 |
| Snuffles/rhinitis | 10 | 16 |
| Fever | 15 | 24.6 |
| Hutchinson's teeth | 1 | 1.65 |

<sup>a</sup> some patients developed more than one clinical feature or had several laboratory findings

As regards clinical findings, the majority of patients, 49 (80.3%), showed bone involvement (Fig 5). Parrot’s pseudoparalysis of the right upper limb was the most common sign. A considerable proportion of patients presented with hepatosplenomegaly (54.1%) as well as dermatological manifestations (50.8%) (Fig 6).

**Fig 5.**
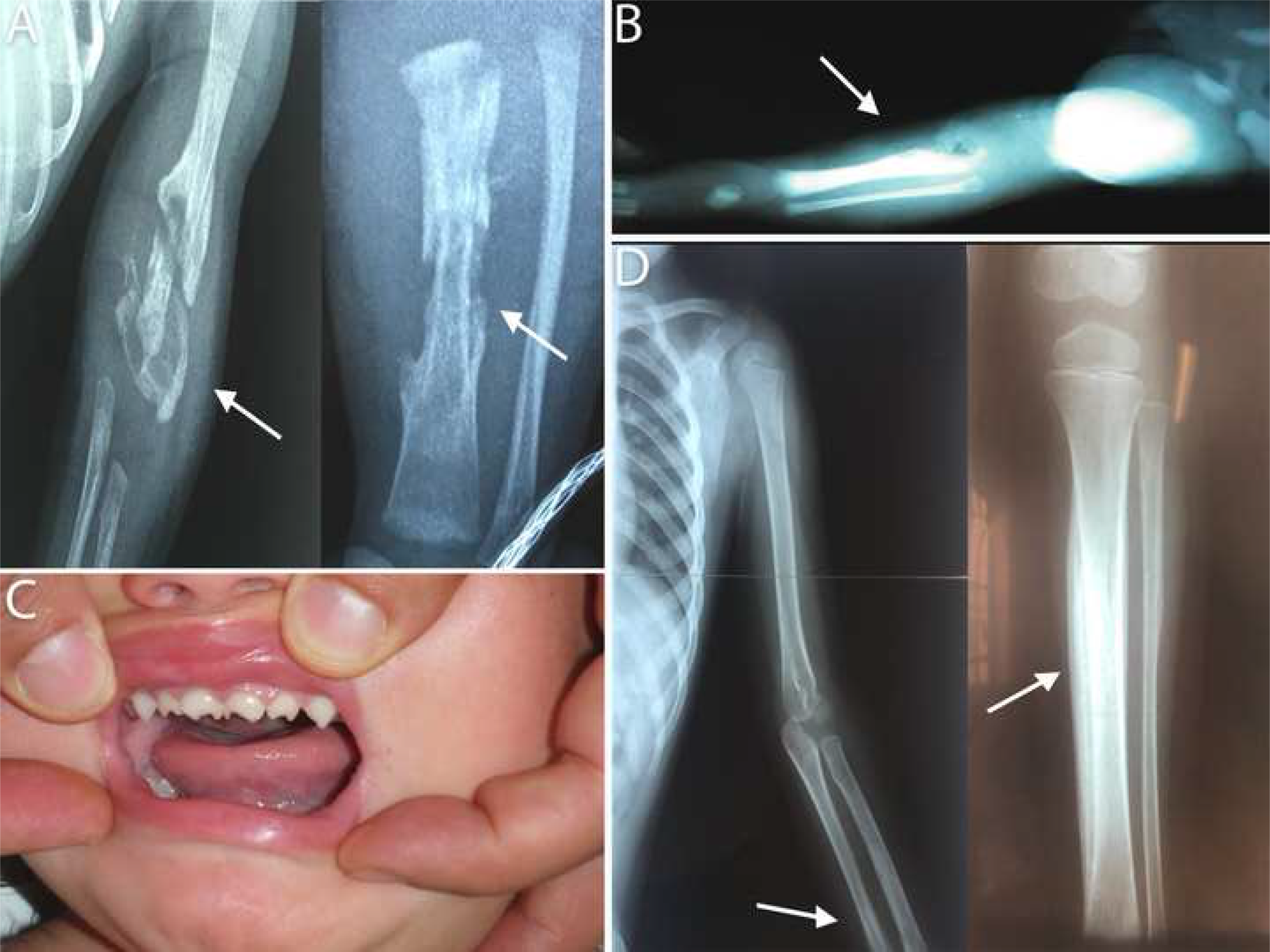
Representative radiological images of bone involvement and Hutchinson’s teeth findings. (A) Anteroposterior view radiographs of a female newborn showing lytic image in the distal region of left tibia and humerus with diffuse periosteal reaction (arrow). (B) Radiographs of a male newborn showing lytic image in the proximal region of right the tibia and fibula. (C) Hutchinson’s teeth in a 2-years old girl (D) Radiographs showing widespread periostitis of the humerus and tibia (arrow) in a 2-years old girl

**Fig 6.**
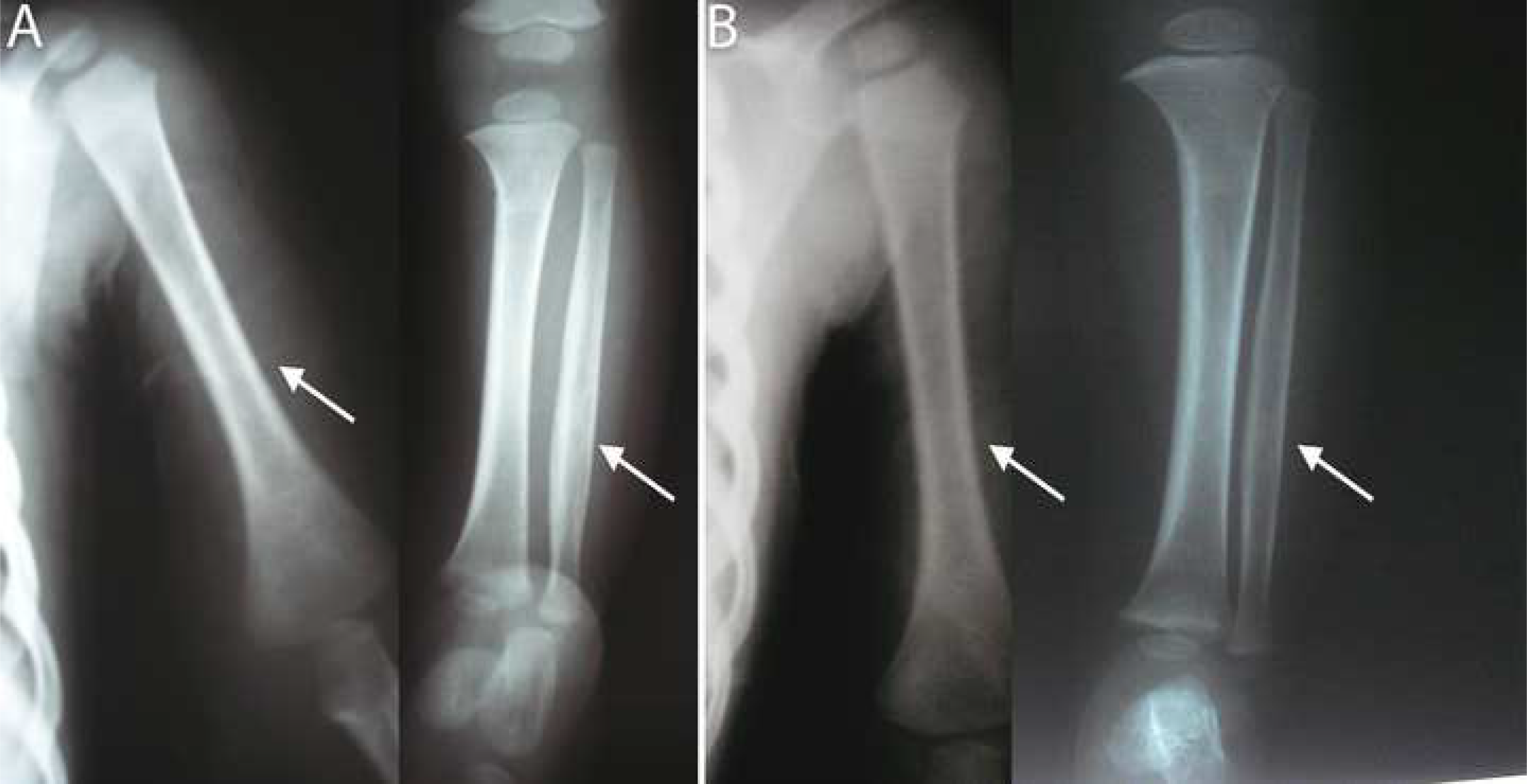
Representative radiological images of the bone lesions evolution after treatment. (A) Anteroposterior view radiographs of a 14 month-old girl showing diffuse periosteal reaction in the left humerus as well as in tibia and fibula (B) Improvement of the lesions after treatment

Complementary studies at diagnosis are presented in Table 3. Alterations in hepatic parameters were observed in 17/47 (36%) of the patients. Severe anemia, with hemoglobin lower than 7.5 g/l and hemodynamic compromise, which required red blood cell transfusion, was observed in 5/61 (8.2 %). Ocular fundus examination showed alterations in 4/36 (2.4%) patients, with retinal pigmentation (n=2) keratitis (n=1) and brightness alteration of the fovea (n=1). Lumbar puncture was performed in 50/61 patients (82%): CSF was altered in 5 (10%) patients, 4 showed reactive VDRL and 1 showed a high white blood cell count. In 9 patients (18%) lumbar puncture was traumatic.

**Table 3.** Complementary studies at diagnosis.

| **Routine laboratory data <sup>a</sup>** | **n** | **%** |
| --- | --- | --- |
| Urinalysis | 45 | 73.7 |
| Normal | 30 | 66.6 |
| Altered | 15 | 33.3 |
| Mild proteinuria | 8 | 17.7 |
| Massive proteinuria | 4 | 8.9 |
| Hematuria | 9 | 20 |
| Leukocyturia | 2 | 4.4 |
| Liver functions | 47 | 77 |
| Normal | 30 | 63.8 |
| Altered | 17 | 36.2 |
| Hyperbilirubinemia. | 14 | 29.7 |
| Alanine transaminase elevation | 12 | 25.5 |
| Complete Blood Count | 51 | 83.6 |
| Anemia | 30 | 58.8 |
| Leukocytosis | 16 | 31.4 |
| Thrombocytopenia | 5 | 9.8 |
| **Other diagnostic' work up studies <sup>a</sup>** | **n** | **%** |
| Ocular fundus examination | 36 | 59 |
| Normal | 32 | 89 |
| Altered | 4 | 11 |
| Keratitis | 1 | 25 |
| Retinal pigmentation | 2 | 50 |
| Fovea alterations | 1 | 25 |
| Bone Radiological Findings | 61 | 100 |
| Normal | 22 | 36 |
| Altered | 36 | 59 |
| Periostitis | 36 | 59 |
| Osteochondritis/osteomyelitis | 6 | 9.8 |
| Fracture | 3 | 4.9 |
| Osteolytic lesions | 2 | 3.2 |
| Missed diagnosis | 3 | 5 |
| Cranial ultrasound | 23 | 37.7 |
| Normal | 23 | 100 |
| Hearing evaluations | 19 | 31.1 |
| Normal | 18 | 94.7 |
| Abnormal | 1 | 5.3 |

<sup>a</sup> some patients developed more than one clinical feature or had several laboratory findings

TPA serology at diagnosis: RPR/VDRL showed median titers of 32 dilution (IQ_25-75_: 4 - 128), and TPHA showed median titers of 640 dilution (IQ_25-75_: 160 - 2560).

As regards comparison of mother and child RPR/VDRL titers, 38 binomial samples were available. A considerable number (39.4%) of patients did not reach 4-fold titers more than their mothers and in 21% the titers were the same or even lower. (Table 4)

**Table 4:** Nontreponemal comparative testing in mothers and child.

| RPR determination/ Matching | 4-fold | 2-fold | Equally | Lower |
| --- | --- | --- | --- | --- |
| Child vs mother's titers | 23 (60.5%) | 4 (10.5%) | 3 (7.9%) | 8 (21%) |

Curiously, in our cohort, one patient (1/61) tested negative for RPR at birth but eventually developed symptoms of late CS (Hutchinson’s teeth) and RPR was positive (Fig 6C).

Treatment: in 60/61 subjects, intravenous aqueous crystalline penicillin G for 10-14 days was prescribed. Doses were adjusted to patients’ age and weight covering possible involvement of CNS, following national guidelines. One patient started treatment with ceftriaxone 50 mg/kg/day because of suspected sepsis. Upon diagnosis of syphilis, 10 days of treatment was completed.

Jarisch-Herxheimer reaction (fever and some vasomotor signs) occurred in 9/61 (14.7%) patients. Its onset was within 12 hours after treatment with a maximum duration of 36 hours.

Overall 47/53 (89%) symptomatic patients recovered after treatment (Fig 7). However, a three-month-old patient with severe anemia, acute kidney failure and sepsis by *Escherichia coli* died. In 4 patients persistent proteinuria was observed as a sequelae of renal involvement and in 1 patient knee arthrosis occurred due to bone damage.

**Fig 7.**
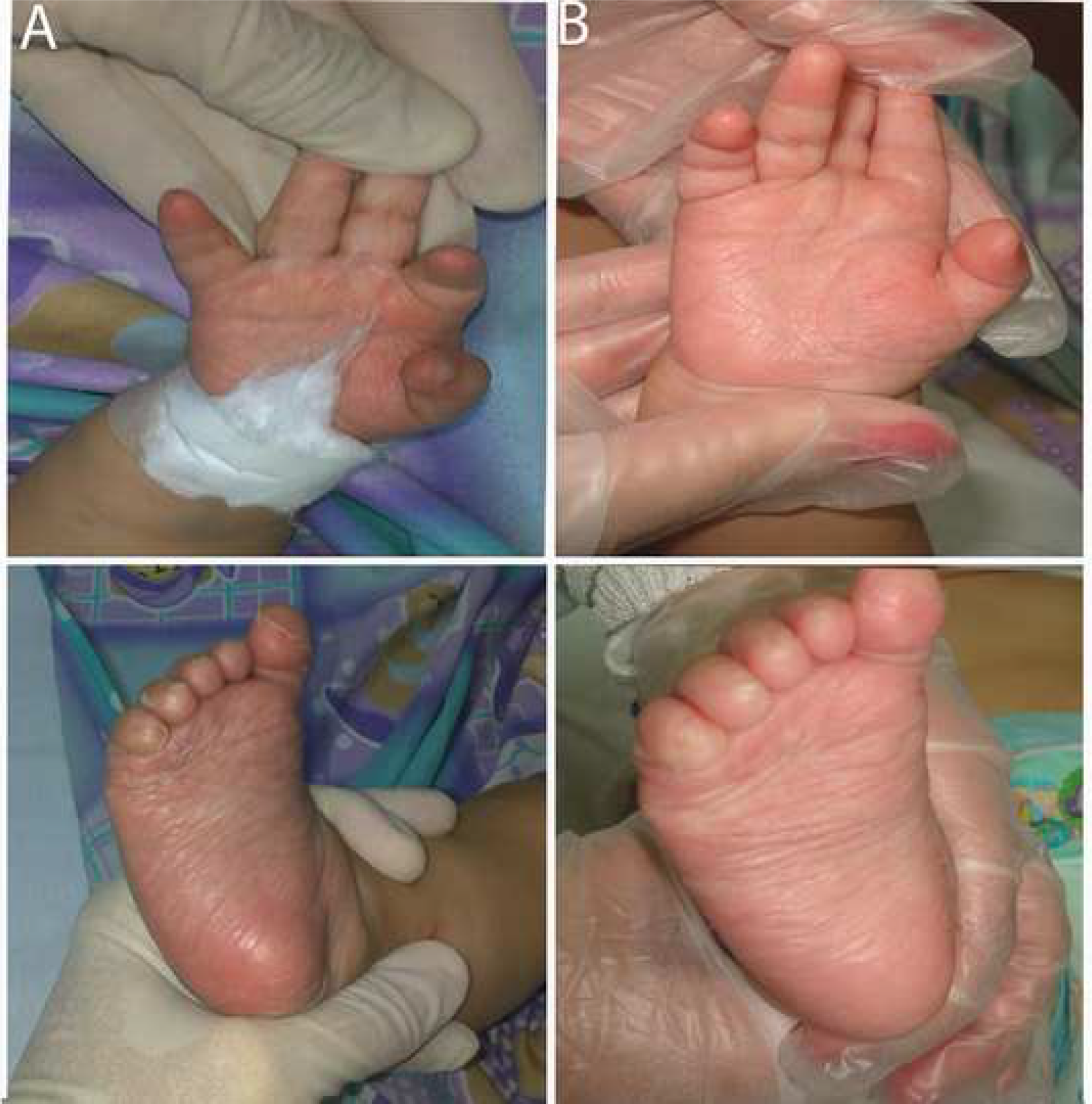
Representative images of dermatological manifestations. (A) Maculopapular rash on the palms and soles in a 4-month-old boy. (B) Improvement of the lesions two days after penicillin treatment in the same patient.

Serological follow-up: Serological data for follow-up analysis was available for 34/61 patients. The media follow-up time was 14.4 months (95% CI: 5.9-23). A decrease of VDRL/RPR titers was observed reaching seroconversion in 31/34 (91%) subjects at a median time of 19.2 months after treatment follow-up. Seroconversion time was analyzed with the Kaplan-Meier test (Fig 8). A decrease in TPHA titers was also observed and only 2 patients showed seroconversion at 5 years of follow-up.

**Fig 8.**
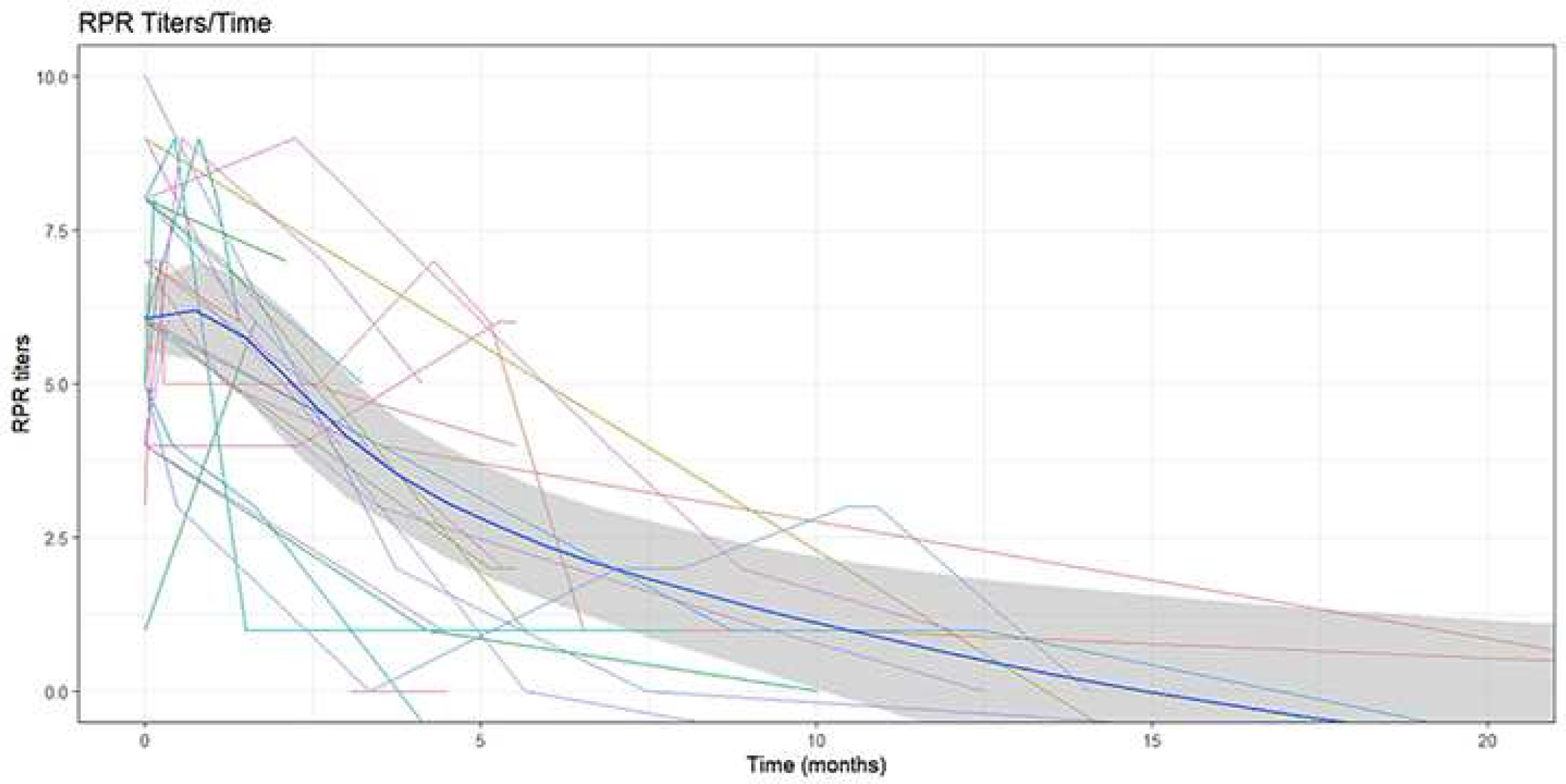
After treatment follow-up RPR titers in 34 congenital syphilis treated patients. Figure shows log titers of RPR changes over time. Dark blue line represents the smooth regression of the data.

## Discussion

Over the last 30 years our service, as a pediatric referral center, assisted over 61 patients suffering from CS, which is a potentially eradicable disease. In Argentina an increase in primary syphilis was reported over the last few years [3]. In order to see whether the rising trend of primary syphilis had an impact on the occurrence of CS, we conducted a retrospective analysis of a cohort of assisted CS patients in our hospital.

Our results showed an increase in the number of cases of CS in recent years. Interestingly a bimodal curve with a peak of cases in the early 90’s was observed and another peak in 2017. A similar trend has been highlighted in several countries [5,6]. The first increase was attributed to the new diagnostic definition of CS in 1988 [5], and the second peak was due to an increase in reported cases of primary syphilis in women from 2015 onwards [6,7], a phenomena that has also occurred worldwide [3].

Despite the fact that 96% of the births took place in medical centers in our country, a high percentage of women were not adequately screened for syphilis during pregnancy [3,8]. This leads to missed opportunities to prevent the transmission of TPA to the fetus [2,9]. In our study, only 47% of pregnant women completed adequate prenatal serological screening and only four mothers were treated. Unfortunately, we do not have sufficient data to rule out a therapeutic failure of these mothers.

The WHO estimate that 27% of CS newborns from untreated mothers showed IUGR, prematurity or are born small for gestational age [4]. In our study a high rate of preterm newborns (24.6%) and low birth weight (24.6%) was observed, as was reported worldwide [10, 11]. A high percentage (73%) of our patients were asymptomatic at birth and developed clinical manifestations of CS within the first 2 years of life. It was estimated that two thirds of untreated asymptomatic newborns would develop CS symptoms in the first 3 to 8 weeks of life and almost all at 3 months of life [4]. A delayed diagnosis was observed in our cohort due to inadequate serological studies during pregnancy, which did not alert health professionals to the presence of CS in asymptomatic newborns.

The analysis of the clinical manifestation demonstrated a high morbidity of a preventable disease. Bone involvement was observed in a great number of patients. Bone involvement (particularly osteochondritis) revealed by radiological tests, was the most frequent finding in our study. This proved to be a useful marker of CS; as suggested by others authors [4,12]. Hepatosplenomegaly has also been reported as one of the most common findings [11]. In our study, both hepatosplenomegaly and dermatological manifestations followed in frequency after bone lesions. The high rate of bone involvement observed may be due to the fact that we assist infants with CS more than newborns, given that we are a referral pediatric center without a maternity ward.

As regards laboratory findings, hepatitis, jaundice and anemia are well-known manifestations of CS. It has been suggested that this is related to the direct action of TPA causing hepatocellular cholestasis (without anatomical alteration of the bile ducts) [13]. Although 54% of our patients presented with hepatomegaly, only 36% presented with alterations in the hepatic panel. In contrast, anemia was present in a high percentage of patients, requiring blood transfusion in 5/61 cases. These findings are similar to those reported in a previous study by our group [8].

In order to evaluate CNS involvement, CSF testing (cell count, protein, and VDRL) has been recommended since neurosyphilis is the manifestation most feared [10,14]. In our study, only 10% (5/50) of patients whose CSF was evaluated showed alterations. However, there are many issues with diagnostic accuracy for example: white-cell count and protein values vary with age; VDRL/RPR antibodies can passively transfer into the CSF, and TPA was detected with normal CSF results.[18].

These facts justified a treatment regimen that covers, in dose and time, CNS involvement as was prescribed in our patients. The need for CSF evaluation has been questioned, due to its lack of accuracy in confirming or discarding CNS involvement [10,15]. It was suggested that asymptomatic newborn lumbar puncture examination be avoided [1] [16]. In our study lumbar puncture was of limited usefulness as a result of blood contamination and the low VDRL sensitivity and did not change the therapeutic regimen in our patients. Also no differences in clinical after-treatment evolution was observed between patients with or without CSF evaluation confirming that lumbar puncture did not provide reliable data as a prognostic marker.

We observed that 89% of asymptomatic patients recovered after treatment with intravenous aqueous crystalline penicillin G. Although 14.7% showed Jarisch-Herxheimer reaction, these events were self-limiting. One patient received daily intravenous ceftriaxone with a favorable clinical evolution. The possibility of using alternative drugs to penicillin for the treatment of syphilis is a field that needs more research. We believe that ceftriaxone is a good alternative given the worldwide shortage of penicillin [17–19].

For the diagnosis of syphilis, a wide range of tests are available but several factors must be considered. Direct identification of TPA by dark field microscopy is not routinely used because it is only useful in symptomatic patients with primary syphilis and has low sensitivity [20]. In CS reactive serological results in newborns and infants results can be confusing due to passive placental transfer of maternal antibodies [20,21]. This often results in unnecessary treatment and hospitalization of asymptomatic newborns with reactive serology from mothers with syphilis, which overloads the health system. Non treponemal antibody titers 4 times higher in newborns than those of the mother has been proposed as a marker of active CS infection [10]. [22]. However in our study, 4-fold higher RPR titers were absent in a considerable proportion of infected patients and did not help to reach CS diagnosis [20].

Non-treponemal tests (VDRL/RPR) detect anti-cardiolipin antibodies. The sensitivity of this test increases with higher titers. A higher concentration of VDRL/RPR antibodies is related to the activity of the infection. In our population, a sustained decrease in antibody titers was observed as an indicator of therapeutic response (Fig 4).

The treponemal (TPHA, FTA-Abs) test detects TPA antibodies and is used to confirm reactive VDRL/PRP. The contribution of the treponemal (TPHA, FTA-Abs) test in the diagnosis of CS was questioned. In our study TPHA titers remained reactive after treatment so this test was not useful for treatment monitoring.

Our results reflect that it is vital to study all asymptomatic newborns whose mothers have syphilis, and to carry out a comprehensive clinical and laboratory evaluation including long bone radiography due to the high incidence of bone compromise observed in CS. If VDRL/RPR becomes negative during follow up, showing the disappearance of passively transferred maternal antibodies, CS diagnosis is ruled out [21]. Maternal-specific treponemal antibodies may persist for up to 12 months [6,10]. If VDRL/RPR remains reactive at 12 months, the infection is confirmed.

New techniques for CS diagnosis are needed since a long term follow-up is required to confirm the diagnosis, especially in asymptomatic infants [2]. Nucleic acid-based amplification assays, such as polymerase chain reaction (PCR) have greatly improved sensitivity and specificity of direct detection of TPA. However, there is sparse information regarding TPA-PCR in clinical settings. A clinical study is under development in our center (Clinical trials Identifier: NCT04084379) in order to validate the sensitivity and specificity of this test for CS diagnosis.

Low patient adherence to follow up was observed. We assist vulnerable and low-income populations with high internal migration, which generates real-world problems in complying with medical follow-up. This constituted a barriers to adequate follow-up in our cohort.

There are many issues that make CS a great challenge for the pediatrician, as is shown in this study. A high number of symptomatic infants with a preventable and a completely curable fetal infection was observed resulting from inadequate detection and treatment of syphilis during pregnancy.

It is mandatory to improve the health care of syphilis, both in terms of screening and treatment, during pregnancy.

## Data Availability

all data are fully available without restriction

## Acknowledgments

The authors wish to thank native speaker Lesley Speakman for revising the english of the manuscript.

## Supporting information

**S1 File. STROBE checklist**.

